# A Reduced Digital Motor Assessment using Multimodal Biomarkers in Parkinson’s Disease

**DOI:** 10.64898/2026.10.05.26364743

**Authors:** Júlia Rey Vilches, Trine Hørmann Thomsen, Bo Biering-Sørensen, Sadasivan Puthusserypady, Silvia Tolu

**Affiliations:** Department of Electrical and Photonics Engineering, Technical University of Denmark, Kongens Lyngby, Denmark; Department of Health Technology, Technical University of Denmark, Kongens Lyngby, Denmark; Department of Neurology, Bispebjerg Hospital, Copenhagen, Denmark; Department of Neurology, Copenhagen University Hospital – Amager and Hvidovre, Hvidovre, Denmark; Department of Public Health, Faculty of Health and Medical Sciences, University of Copenhagen, Denmark

**Keywords:** Parkinson disease, digital biomarkers, surface electromyography, wearable sensors, musculoskeletal modeling, machine learning, motor assessment

## Abstract

**Background:** Wearable sensing and machine learning can support objective assessment of Parkinson’s disease (PD), but performance depends on sensor placement, modality, and analysis, and studies often target selected motor tasks, whereas clinical assessment integrates multiple standardized movements. How discriminative information is distributed across tasks and modalities remains unclear, limiting comparability and translation.

**Objectives:** To quantify standardized Movement Disorder Society Unified Parkinson’s Disease Rating Scale Part III (MDS-UPDRS III) tasks across complementary modalities, determine task- and modality-specific contributions to discrimination, and identify complementary, stable candidate digital biomarkers.

**Methods:** Thirty-one participants with PD (Hoehn and Yahr stages I–III) and 30 age- and sex-matched healthy controls performed seven MDS-UPDRS III tasks (items 3.4–3.8, 3.10, 3.15). Unilateral surface electromyography from task-specific muscles and full-body inertial motion capture were acquired, which drove subject-specific musculoskeletal simulations. A leakage-free classification pipeline (in-fold feature selection, repeated cross-validation, permutation testing) was applied by task and modality, followed by sparse task fusion and stability-based biomarker selection.

**Results:** Discrimination was concentrated in four upper-limb tasks, with pronation–supination (3.6) performing best (ROC-AUC 0.94; balanced accuracy 0.92). Multitask multimodal fusion maintained discrimination (ROC-AUC 0.93; balanced accuracy 0.92; sensitivity 0.87; specificity 0.97), retaining finger tapping (3.4), hand movements (3.5), pronation–supination (3.6), and postural tremor (3.15). Eighteen stable candidate biomarkers captured complementary alterations in movement execution, muscle activation, coordination, tremor, and proximal stabilization.

**Conclusions:** In this cohort, discriminative information converged on a four-task upper-limb assessment and a physiologically interpretable multimodal biomarker profile. These findings support shorter, clinically anchored digital motor assessment, pending independent clinical validation.

## 1 Introduction

The diagnosis of Parkinson’s disease (PD) remains clinical, and objective quantification of motor dysfunction has become a major focus of PD research, particularly when motor symptoms remain mild [1–3].

The Movement Disorder Society–Unified Parkinson’s Disease Rating Scale Part III (MDS-UPDRS III) provides a standardized battery of motor tasks widely used for clinical motor assessment and monitoring [4]. Each item, however, is scored by visual observation on a coarse ordinal scale, with recognized limitations in inter-rater reliability and sensitivity to subtle change [1, 5–8]. Objective quantification of these tasks could provide continuous, sensitive measures while preserving their established clinical relevance and comparability across studies.

Wearable sensing combined with machine learning (ML) has repeatedly distinguished PD from healthy aging with high accuracy [9, 10], yet clinical translation remains limited [11]. Studies vary considerably in the motor tasks investigated, sensing modalities employed, features extracted, and validation strategies used [9, 10]. This methodological heterogeneity limits direct comparison of performance outcomes across cohorts and studies [12–14].

Analytical practice compounds this heterogeneity [15]: models are frequently tuned and the best-performing feature subsets reported within a single dataset, without evaluating their stability across resampling [16, 17], while leakage between feature selection and validation is common [18]. Increasing model complexity can additionally reduce physiological interpretability and clinical applicability [19, 20]. The relevant question is therefore not only whether mild PD can be discriminated from controls, but which measurements are sufficiently robust, interpretable, and clinically anchored to support translation.

A streamlined digital motor assessment is ultimately desirable, but meaningful simplification requires understanding the complementary contribution of standardized motor tasks and sensing modalities, neither of which is established. Most studies examine a single task or study-specific movement and report its accuracy in isolation [13, 21], leaving unclear which tasks provide complementary information and which might be redundant, and thus whether protocols can be shortened without losing clinically relevant information [13, 22].

Single-modality designs further constrain interpretation [23]. Surface electromyography (sEMG) characterizes neuromuscular activation, inertial measurement unit (IMU) sensing quantifies movement kinematics, and IMU-driven subject-specific musculoskeletal (MSK) simulation estimates joint and muscle-level biomechanics [24]. Integrating these measurement sources may therefore improve interpretation of discriminative features in relation to PD motor symptoms.

In this study, seven standardized MDS-UPDRS III motor tasks were quantified using unilateral sEMG and IMU-driven subject-specific MSK simulation in individuals with PD and age- and sex-matched controls. An interpretable [19], leakage-free analytical framework was applied to determine (1) which tasks and measurement modalities discriminate PD from controls, (2) whether integrating information across tasks improves discrimination, (3) which tasks provide complementary rather than redundant information, and (4) which stable features constitute interpretable candidate digital biomarkers.

## 2 Methods

### 2.1 Participants

This single-center, cross-sectional controlled study was conducted at the Movement Disorder Clinic at Rigshospitalet Glostrup, Denmark, with participants recruited between November 2024 and April 2025. Individuals with idiopathic PD were recruited through the clinic and were eligible if they were in Hoehn and Yahr (HY) stages I–III [25] and able to stand and walk independently. Healthy controls (HC) were recruited as age- and sex-matched volunteers.

Exclusion criteria included cognitive impairment that could affect study compliance (Montreal Cognitive Assessment score < 22 [26]), other neurological or musculoskeletal disorders affecting motor function, and orthopedic injury or surgery affecting mobility. Participants with PD were assessed under usual clinical conditions without medication withdrawal. Further protocol and recruitment details are reported elsewhere [27].

The Regional Health Research Ethics Committees in the Capital Region waived the need for ethical approval (no. H-24052002). The study was conducted in accordance with the Declaration of Helsinki, and all participants provided written informed consent.

### 2.2 Data Sources and Study Procedures

Wearable sensor data were collected during a single in-clinic assessment, following the previously reported protocol [27]. For participants with PD, time since last dopaminergic medication dose was recorded, and the more-affected side was identified by patient report. Participants were fitted with two types of wearables, recorded synchronously: a 17-sensor wireless IMU motion-capture suit (MVN Awinda, Movella) for whole-body kinematics, and wireless sEMG sensors (MuscleBAN; bipolar Ag/AgCl electrodes, 1000 Hz) over task-relevant muscles on the reported more-affected side for PD, and dominant side in HC. Electrode placement and muscles are provided in Supplementary Table S1.

Seven standardized MDS-UPDRS III tasks were selected to represent clinically relevant upper- and lower-limb motor function [4, 27]: finger tapping (T1, item 3.4), hand movements (opening and closing, T2, item 3.5), wrist pronation– supination (T3, item 3.6), postural tremor of the hands (arm hold, T4, item 3.15), toe tapping (T5, item 3.7), gait including a 10-m walk-and-turn (T6, item 3.10), and leg agility (foot stomping, T7, item 3.8).

Participant anthropometric measurements were used to calibrate the IMU motion-capture model. The whole-body kinematics drove a subject-specific MSK model in the AnyBody Modeling System [28]. Body-segment lengths were scaled from the calibrated motion-capture model, while measured body mass was provided as a model input. Simulations estimated subject-specific biomechanical variables not directly accessible from IMU recordings. Together with the recorded sEMG signals, these data formed the basis for multimodal feature extraction, following the framework proposed previously [24].

### 2.3 Signal Preprocessing and Feature Extraction

Features were extracted independently for each motor task and sensing modality. Complete signal processing details and feature definitions are provided in Supplementary Methods and Tables S2–S4.

#### 2.3.1 sEMG Features

sEMG signals were band-pass filtered (1–450 Hz, fourth-order Butterworth) and notch filtered at 50 Hz; full-wave rectified and linear envelope (10 Hz low-pass) representations were then derived. Repetitive (T1, T2, T3, T5, T7) and gait (T6) tasks were amplitude-normalized to the 95th percentile of the envelope, applied consistently to the filtered, rectified, and envelope signals. Activation bursts were detected from the envelope, and burst-level features were summarized across repetitions to characterize their central tendency and variability. Postural tremor (T4) was analyzed as a continuous epoch in absolute units (mV). Features characterized amplitude, frequency, timing and rhythm, complexity, and inter-muscle coupling (Supplementary Table S2).

#### 2.3.2 IMU-driven MSK Features

Biomechanical features were extracted from MSK simulations driven by each participant’s whole-body IMU kinematics. Features included muscle activations and forces, joint moments and reaction forces, antagonist co-contraction, and bilateral asymmetry. In addition, simulation-derived kinematic descriptors (joint angles, range of motion, angular velocities, end-point kinematics, gait speed, and positions) were included. Depending on the task, 154–468 features were extracted (Supplementary Tables S3 and S4).

### 2.4 Analytical Framework

#### 2.4.1 Classification Pipeline and Validation

The extracted features were analyzed separately for each motor task using three configurations: sEMG, MSK-derived, and multimodal configuration combining both feature sets. For each task–configuration combination, feature selection and model training were performed exclusively within each cross-validation training fold. Features underwent median imputation, near-zero-variance and correlation filtering (|*r*| *>* 0.80), followed by mutual-information (MI) ranking. The MI-curve knee point determined the candidate feature pool entering *l*_1_-penalized (LASSO) selection, yielding a subset of 3–5 features used for classification. In the multimodal configuration, filtering, MI ranking, and candidate-pool determination were performed separately for sEMG and MSK before concatenation and LASSO selection, preventing the larger feature set from dominating selection.

The selected features were used to train eleven classifiers: logistic regression (LR), linear, polynomial and radial-basis-function support vector machines (SVMs), naive Bayes (NB), *k*-nearest neighbours (kNN), decision tree (DT), random forest (RF), gradient boosting (GB), extreme gradient boosting (XGBoost), and linear discriminant analysis (LDA). The model with the highest cross-validated balanced accuracy (BA) was retained for each task–configuration combination; hyperparameters were not tuned.

Performance was estimated using stratified 10-fold cross-validation. BA, ROC-AUC, sensitivity, and specificity were calculated from the pooled cross-validated predictions. To assess the robustness, cross-validation was repeated over 30 random partitions, and the variability was summarized as 95% intervals across repetitions. Statistical significance was assessed using a 500-label permutation test to verify that classification performance exceeded chance expectations. Feature-selection stability was quantified as the proportion of cross-validation folds in which each feature was retained [17], thereby prioritizing consistently selected candidate digital biomarkers. Implementation details are in the Supplementary Methods.

#### 2.4.2 Modality and Task Fusion

To evaluate the combined information across tasks, cross-validated task-specific PD probabilities were integrated using a task-level fusion model. For each motor task, the cross-validated PD probability from its best-performing classifier was used as input to an *l*_1_-penalized logistic-regression meta-model (LASSO), which integrated task-level predictions into a single subject-level prediction (Fig. 2A). Only cross-validated task probabilities were used as meta-model inputs, ensuring that each participant’s probability was generated by a classifier not trained on that participant; the meta-model was then evaluated using a separate stratified 10-fold cross-validation.

Three fusion models combined sEMG, MSK, and multimodal task-specific predictions. Fused models were evaluated with the same cross-validated metrics as the task-specific classifiers.

#### 2.4.3 Task Complementarity

Task contributions were quantified from LASSO meta-model coefficients, with non-zero coefficients identifying tasks retained in the fused prediction. Selection frequency across repetitions assessed the stability of task retention.

Redundancy was evaluated using pairwise Pearson correlation and mutual information (MI) between cross-validated task-specific PD probabilities. Together, task retention and redundancy were used to identify a candidate reduced set of tasks contributing complementary discriminative information.

#### 2.4.4 Stable-Feature Identification and Biomarker Characterization

Within the tasks retained by the fusion analysis, selection stability was quantified within each feature’s own modality. Features selected in more than 50% of cross-validation folds were considered stable and were retained as candidate biomarkers if they also differed significantly between groups (Mann–Whitney *U* test, *p* < 0.05). Retained features were further characterized in the full cohort by their effect size (Cohen’s *d*) and MI with group status, and principal component analysis (PCA) was used to visualize their joint distribution (Fig. 3C). Features were further characterized by modality, anatomical origin, and plausible relation to PD motor dysfunction.

#### 2.4.5 Sensitivity Analyses

Within the PD group, the multimodal fused PD probability was tested for associations with HY stage and time since last medication dose using Spearman correlations. Associations of the multimodal fused PD probability and age and body mass index (BMI) were assessed using Spearman correlation, and sex differences were assessed using Mann–Whitney *U* tests within each group.

## 3 Results

### 3.1 Participants

The study cohort included 61 participants: 31 individuals with PD and 30 age- and sex-matched HC (Table 1). The groups did not differ significantly in age or sex. Participants with PD were assessed in their usual clinical condition without medication withdrawal. The PD cohort was predominantly mildly affected according to HY stage, while time since diagnosis was heterogeneous, with approximately half of the participants diagnosed within the previous 5 years.

**Table 1:** Participant characteristics.

| Characteristic | HC ($n = 30$) | PD ($n = 31$) | $p$ |
| --- | --- | --- | --- |
| Sex, F/M | 18/12 | 14/17 | 0.37 |
| Age, years | $59.3 \pm 13.5$ [40–79] | $62.6 \pm 9.1$ [43–79] | 0.26 |
| *PD-specific* |  |  |  |
| HY stage, median (IQR) [range] | — | 1 (1.0–2.0) [1–3] | — |
| stage 1 / 1.5 / 2 / 3, $n$ | — | 17 / 4 / 7 / 3 | — |
| Time since diagnosis, years | — | $4.6 \pm 3.2$ [0.02–12] | — |
| < 5 years / $\geq 5$ years, $n$ | — | 15 / 16 | — |
| Time since last dose, h | — | $2.9 \pm 1.9$ [0.25–6.5] | — |

Values are mean ± SD [range] or $n$ , unless otherwise indicated. HY stage is reported as median (IQR) [range]. $p$ -values compare groups using a two-sample $t$ -test for age and a $\chi^2$ test for sex. Patients were evaluated in their usual ON-medication state. Abbreviations: HC, healthy controls; PD, Parkinson's disease; HY, Hoehn and Yahr.

### 3.2 Performance of Standardized Motor Tasks

Discriminative performance varied markedly across tasks, with discrimination concentrated in the four upper-limb assessments (T1–T4; Fig. 1). Pronation–supination (T3) achieved the highest overall performance (multimodal ROC-AUC = 0.94, balanced accuracy = 0.92). Lower-limb tasks showed only low-to-moderate discrimination (ROC-AUC = 0.52–0.72). All four upper-limb tasks performed significantly above chance across all three feature configurations, whereas no lower-limb task was significant in more than one configuration.

**Figure 1:**
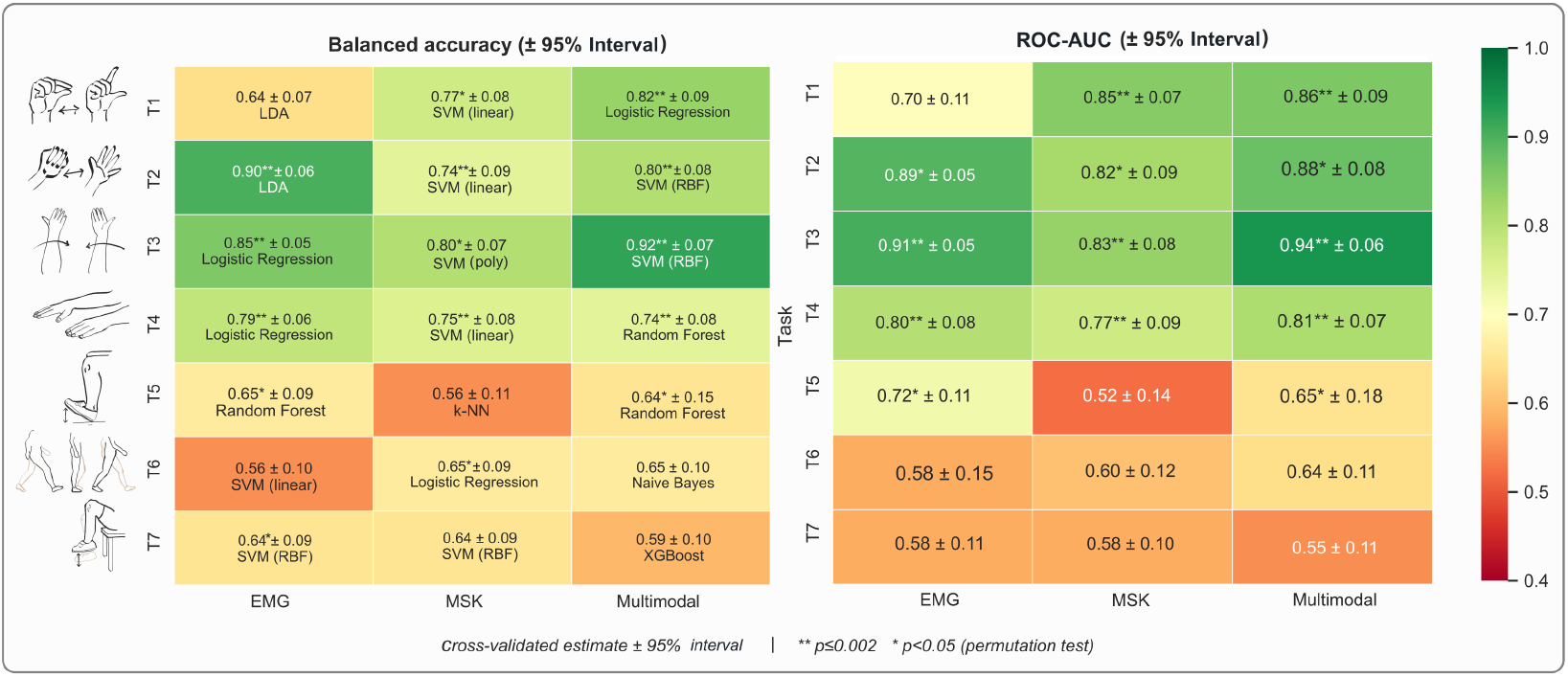
Single-task classification performance. Balanced accuracy and ROC-AUC across seven MDS-UPDRS motor tasks using sEMG, musculoskeletal (MSK), and multimodal features. Values show cross-validated estimates and variability (95% intervals across 30 repetitions); asterisks indicate permutation-test significance.

The performance of the sEMG, MSK, and multimodal configurations was task-dependent. MSK features outperformed sEMG for finger tapping (ROC-AUC 0.85 vs. 0.70), whereas sEMG performed best for hand movements (0.89 vs. 0.82). Multimodal features provided the highest ROC-AUC for pronation–supination (0.94) and postural tremor (0.81), but did not consistently improve performance in the lower-limb tasks.

### 3.3 Task-level Fusion

The multimodal fused model achieved balanced accuracy of 0.92, sensitivity of 0.87, specificity of 0.97, and ROC-AUC of 0.93 (Fig. 2B), compared with 0.89/0.87/0.90/0.92 for sEMG fusion and 0.84/0.81/0.87/0.91 for MSK fusion. It therefore maintained the high discrimination achieved by the strongest individual task (T3 multimodal: balanced accuracy = 0.92; ROC-AUC = 0.94), while integrating information across tasks and increasing specificity slightly from 0.95 (Supplementary Table S5) to 0.97 (Fig. 2B).

**Figure 2:**
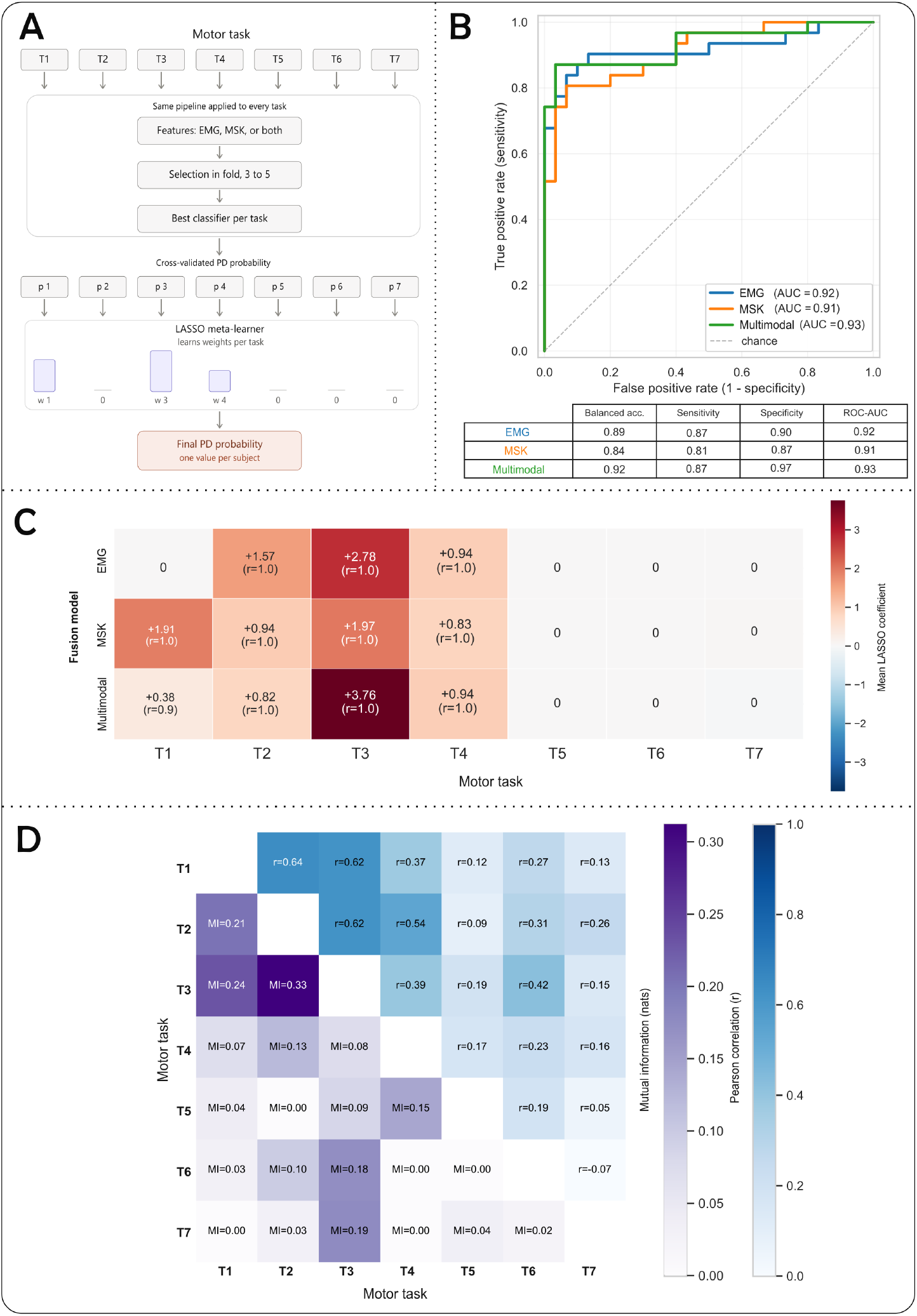
Task fusion and complementarity. (A) Task-level fusion framework. (B) Performance of sEMG, MSK, and multimodal fusion. (C) Task contributions and selection frequency. (D) Pearson correlation and mutual information between task-specific cross-validated prediction probabilities.

### 3.4 Task Complementarity

In the multimodal fusion model, T2, T3, and T4 were retained in 100% of repetitions and T1 in 90%, whereas T5–T7 were consistently assigned zero coefficients (Fig. 2C; Supplementary Table S6). Pronation–supination contributed the largest mean coefficient (+3.76).

Retained tasks differed across the three fusion models. The sEMG model retained T2–T4, whereas the MSK model retained all four upper-limb tasks, with T1 and T3 among the strongest contributors (Fig. 2C).

The three repetitive upper-limb tasks showed the greatest correlations (Pearson *r* = 0.62–0.64; MI = 0.21–0.33), whereas postural tremor (T4) showed lower agreement with them (*r* = 0.37–0.54; MI = 0.07–0.13; Fig. 2D), indicating a less redundant prediction pattern than the repetitive upper-limb tasks. Together with its consistent retention in the fusion model, this supported a complementary contribution of the postural tremor assessment. These analyses supported a candidate reduced assessment comprising the four upper-limb tasks.

### 3.5 Stable Candidate Digital Biomarkers

Eighteen stable candidate digital biomarkers were identified across the four retained upper-limb tasks (Fig. 3A). Ten were derived from sEMG and eight from MSK, spanning complementary neuromuscular and simulation-derived biomechanical features localized to the recorded upper limb (Fig. 3B; Supplementary Table S8).

**Figure 3:**
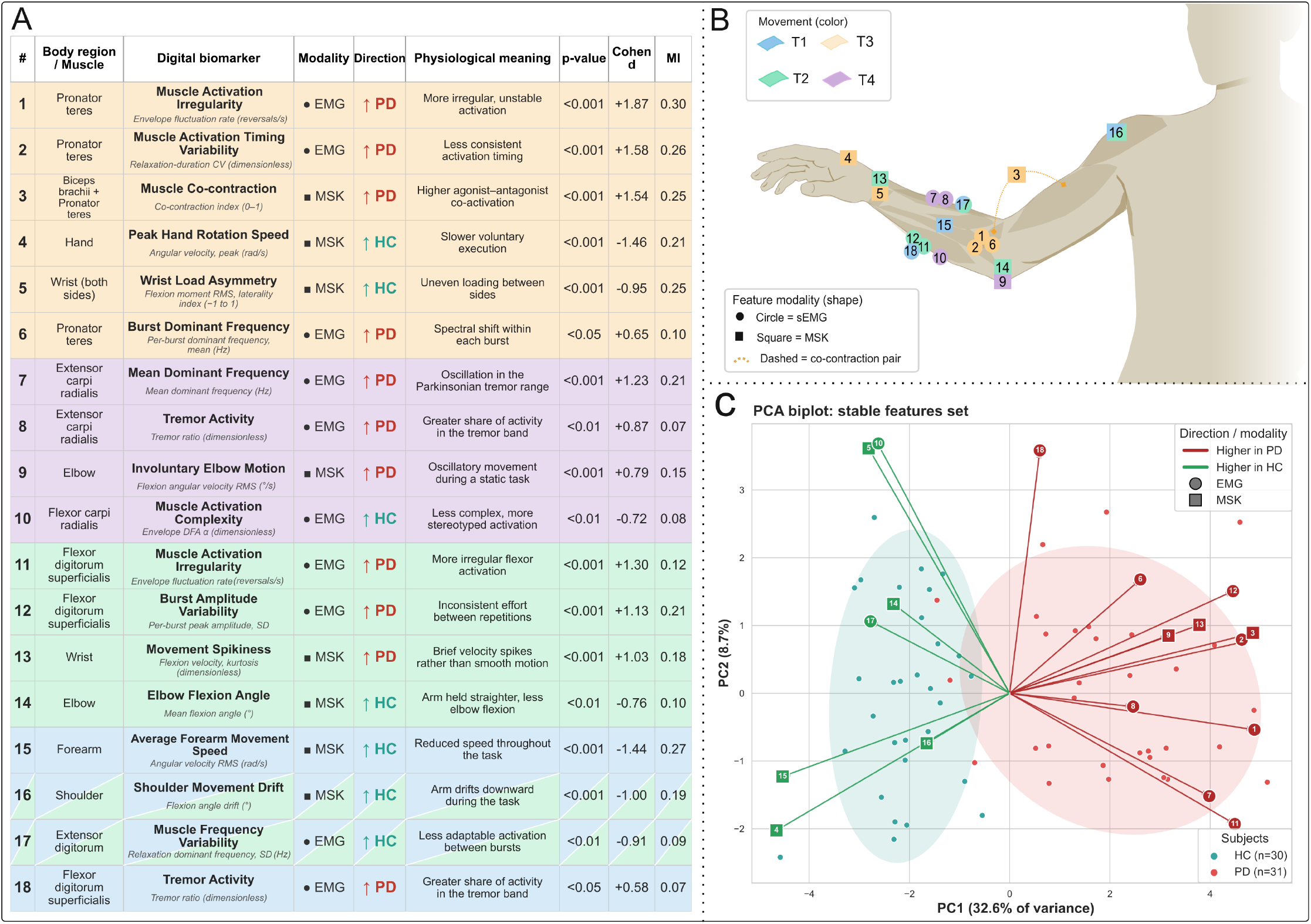
Stable candidate digital biomarkers. (A) Eighteen stable biomarkers across retained tasks. (B) Anatomical distribution by task and modality. (C) Principal component analysis of the stable biomarker set, with 95% intervals and feature loadings.

The largest between-group effects were observed during pronation–supination, including pronator teres activation irregularity (Cohen’s *d* = +1.87), burst-timing variability (+1.58), biceps–pronator teres co-contraction (+1.54), and reduced peak hand rotation speed (−1.46). Additional large effects included flexor digitorum superficialis activation irregularity during hand movements (+1.30) and reduced forearm movement speed during finger tapping (−1.44) (Supplementary Fig. S2).

Across modalities, the retained features captured complementary abnormalities in movement speed and amplitude, agonist–antagonist co-activation, tremor-related activity, and muscle activation regularity. Principal component analysis of the 18-feature set showed separation between groups along the first component, which explained 32.6% of the variance (Fig. 3C, Supplementary Fig. S1).

These findings identify a compact set of physiologically interpretable measurements, distributed across four standardized motor tasks and spanning cardinal domains of PD motor dysfunction.

### 3.6 Sensitivity Analyses

Within the PD group, the multimodal fused PD probability was not associated with HY stage (*ρ* = +0.10, *p* = 0.58) or time since last medication dose (*ρ* = +0.25, *p* = 0.17). The analysis of HY stage had limited resolution because 21 of 31 participants were stages 1–1.5. Age, sex and BMI were not associated with the fused probability in either group (Supplementary Table S7).

## 4 Discussion

This study evaluated seven standardized MDS-UPDRS III motor tasks using sEMG and IMU-driven subject-specific MSK simulation. Discriminative information was concentrated in four upper-limb tasks, with task and measurement-source complementarity supporting a candidate reduced assessment that maintained high PD–HC discrimination. Across these tasks, 18 stable candidate digital biomarkers captured complementary abnormalities in muscle activation, movement kinematics, agonist–antagonist coordination, and tremor-related activity.

### 4.1 Task Complementarity and Protocol Reduction

The concentration of discrimination in the upper limb is consistent with motor features commonly observed in mild PD. Motor signs commonly emerge asymmetrically and distally [29], whereas axial and postural involvement becomes more prominent with disease progression [30]; prominent early postural instability is also considered a warning feature for atypical parkinsonism [31]. Consistent with this pattern, all four upper-limb tasks showed discrimination, whereas toe tapping, gait, and leg agility contributed little to the sparse multimodal fusion. This should not be interpreted as evidence that lower-limb assessment is clinically uninformative; rather, the lower-limb tasks provided limited additional discriminative information in this predominantly mildly affected cohort.

Importantly, pronation–supination alone reached discriminative performance comparable to the fused model, indicating that fusion did not improve peak discrimination. However, fusion combined partially redundant information across motor behaviours, with postural tremor contributing a more distinct prediction than the three repetitive upper-limb tasks. Combining tasks may therefore provide a broader representation of motor dysfunction than relying on a single movement, particularly in a heterogeneous disorder such as PD, where main motor symptoms may vary between individuals [3].

The same complementarity was observed across measurement sources. MSK features were particularly informative during finger tapping, sEMG during hand movements, and multimodal representations during pronation–supination and postural tremor. Thus, no measurement source consistently provided the best representation across tasks.

The preferential contribution of upper-limb tasks may reflect greater sensitivity of distal repetitive movements to subtle bradykinetic and asymmetric motor abnormalities in this cohort, although this remains hypothesis-generating because task-specific clinical severity was not examined. This task dependence may help explain the variable sensor configurations used across digital PD studies and suggests that sensor selection should be considered together with the movement [12, 15, 22].

Together, task and measurement-source complementarity provide a basis for reducing the assessment: preserving a small set of standardized movements that sample complementary motor information. All four retained tasks correspond to MDS-UPDRS III items and can be performed seated, supporting their further evaluation as a short, clinically anchored protocol [27]. The configuration remains a candidate reduced assessment rather than a replacement for the complete motor examination.

### 4.2 Physiological Interpretation of Candidate Biomarkers

The 18 stable candidate digital biomarkers provide a previously unreported combination of physiological and biomechanical measurements describing complementary aspects of motor dysfunction. Participants with PD showed reduced hand and forearm movement velocities and amplitude, greater irregularity and variability in forearm muscle activation, increased antagonist co-contraction, tremor-related activity and postural stabilization while performing predominantly distal tasks. The largest effects occurred during pronation–supination, including irregular pronator teres activation, variable relaxation timing, increased biceps–pronator teres co-contraction, and reduced hand rotation speed. During postural tremor, abnormalities were detectable not only in sEMG tremor-band activity but also as involuntary elbow motion. These measurements therefore quantify recognizable manifestations of PD motor dysfunction rather than functioning solely as classifier features.

Some alterations were recovered independently during more than one standardized task. Shoulder flexion-angle drift was retained during both finger tapping and hand movements, as was variability in the dominant frequency of extensor digitorum activity during relaxation periods. Their recurrence across distal repetitive tasks suggests characteristics of upper-limb control rather than isolated task-specific differences. Shoulder flexion drift indicates that discrimination during distal tasks also emerged in proximal limb behaviour. This may reflect altered stabilization, compensatory movement, or proximal motor control, although the underlying mechanism cannot be distinguished here.

The biomarker set also included established signatures of bradykinesia and hypokinesia, consistent with associations of reduced movement amplitude with hypokinesia, velocity with bradykinesia, movement variability with interruptions in motor outputs, and frequency-domain measures with tremor [22]. The present findings extend that framework by showing that these abnormalities can coexist and recur across standardized tasks, rather than representing isolated task-specific markers. Thus, PD-associated motor dysfunction may be reflected not only in how far or how fast a movement is executed, but also in its consistency, antagonist coordination, and proximal limb behaviour.

### 4.3 Contribution of Subject-Specific MSK Simulations

Many simulation-derived biomarkers were kinematic or spatiotemporal measures that could be derived directly from IMU sensing. The added value of subject-specific MSK simulation was not simply the generation of more kinematic descriptors. Rather, it related measured whole-body movement to joint moments and reaction forces, ground reaction forces, muscle forces, and coordination between muscles not instrumented with sEMG [24].

Antagonist co-contraction illustrates this contribution. Increased co-contraction has been reported in PD and associated with mechanisms relevant to rigidity [32, 33]. Simulation allowed antagonist coordination to be examined beyond the two muscles instrumented with sEMG, including deeper or non-instrumented muscle pairs. However, these estimates should not be interpreted as direct measurements of rigidity, but as candidate biomechanical correlates of altered agonist–antagonist coordination during voluntary movement.

Although gait-related discrimination was limited in this cohort, simulation-derived biomechanical measures may become more informative with greater gait and postural dysfunction and warrant evaluation across broader disease stages and motor phenotypes [14, 31, 34, 35].

### 4.4 Effect of PD Medication on the Assessment

Dopaminergic treatment can improve movement amplitude and velocity, although these measures are not fully normalized [36]. Importantly, the sequence effect–the progressive decrement or increasing irregularity of movement amplitude and velocity across repeated actions–may persist under treatment [37, 38]. This may help explain why measures of movement consistency remained informative under treatment when speed or amplitude are partially restored.

Several of the candidate biomarkers characterized the consistency of motor output rather than its mean magnitude: activation fluctuation, burst-to-burst variability, relaxation-frequency variability, and complexity measures were retained alongside reductions in angular velocity and movement amplitude. This remains hypothesis-generating as neither the sequence effect nor medication effects were directly tested.

Medication exposure was not standardized because participants were assessed under their usual treatment conditions without medication withdrawal. The multimodal fused prediction was not associated with time since last medication dose, but this does not establish medication independence. Given variation in ON/OFF definitions across PD studies [15, 39], direct within-participant ON/OFF assessment is required to determine which candidate biomarkers are treatment-sensitive and which persist across medication states.

### 4.5 Analytical Robustness and Candidate Biomarker Status

The framework aimed not to maximize classification performance, but to identify features that remained informative across repeated resampling and were interpretable by design rather than through post hoc explanation of the classifier. Each candidate is specified by its task, anatomical origin, computational definition, and plausible relation to PD motor dysfunction, providing predefined targets for subsequent studies. This is important if data-driven features are to progress from cohort-specific predictors toward clinically interpretable digital measures [11, 15].

### 4.6 Limitations and Generalizability

The cohort was relatively small and recruited at a single center, limiting generalizability. The concentration of discriminative information in upper-limb tasks should therefore be interpreted within the characteristics of this predominantly mildly affected cohort and may differ in populations with more pronounced lower-limb, gait, or postural involvement.

sEMG recordings were unilateral and restricted to task-specific muscles, potentially missing bilateral abnormalities or information from non-instrumented muscles. MSK simulations extend biomechanical coverage beyond the instrumented muscles; however, simulation-derived variables are model-based estimates and depend on model scaling, muscle recruitment assumptions, and the accuracy of measured kinematics.

Finally, repeated cross-validation and feature-selection stability provide internal validation rather than independent replication. The proposed four-task assessment and 18 candidate digital biomarkers were identified within the same cohort and should therefore be considered preliminary until their direction, magnitude, discriminative contribution, and stability are replicated in larger, independent, preferably multicenter cohorts.

### 4.7 Conclusion

In this predominantly mildly affected cohort, discriminative information was concentrated in four upper-limb tasks, from which 18 stable candidate digital biomarkers captured abnormalities in movement execution, muscle activation and variability, antagonist coordination, and postural tremor. The identified biomarkers, characterized by their task, measurement source, anatomical origin, and computational definition, provide specific targets whose direction, magnitude, and discriminative value can be directly tested in independent cohorts. Their combination across complementary tasks and measurement sources enabled a candidate four-task assessment that retained high PD–HC discrimination while reducing assessment complexity. If independently validated, this approach could complement the neurological examination by providing objective quantitative measures of PD-associated motor abnormalities from a short set of familiar MDS-UPDRS III tasks.

## Supporting information

Supplementary material including more detailed methods, and supplementary tables and figures

## Acknowledgements

We would like to thank all participants for their contribution to this research and the personnel at the Movement Disorder Clinic, Rigshospitalet Glostrup, for their support throughout the study. We particularly thank Nikolaj la Cour Karottki for his contributions to shaping the study protocol and supporting participant recruitment.

## Author’s Roles

(1) Research Project: A. Conception, B. Design, C. Data Collection, D. Data Analysis; (2) Statistical Analysis: A. Design, B. Execution, C. Review and Critique; (3) Manuscript Preparation: A. Writing of the First Draft, B. Review and Editing, C. Approval of Final Version.

J.R.V: 1A, 1B, 1C, 1D, 2A, 2B, 3A, 3B, 3C

THT: 1A, 1B, 1C, 3B, 3C

BBS: 1A, 1B, 1C, 3B, 3C

SP: 1A, 1B, 2A, 2C, 3B, 3C

ST: 1A, 1B, 2A, 2C, 3B, 3C

## Financial Disclosures

The authors declare that there are no disclosures to report.

## Data Availability Statement

The data that support the findings of this study are available from the corresponding author upon reasonable request and subject to applicable ethical, privacy, and institutional data-sharing requirements. The data are not publicly available due to privacy and ethical restrictions. Additional data and methodological information supporting the analyses and findings are provided in the Supplementary Material.

## Notes

### Competing Interest Statement

The authors have declared no competing interest.

### Clinical Protocols

https://ieeexplore.ieee.org/abstract/document/11253014

### Author Declarations

The Regional Health Research Ethics Committees in the Capital Region in Denmark waived the need for ethical approval of the project (no. H-24052002).

### Funding

- Discovery Grant DTU Skylab
- DTU Alliance PhD Fund

