## Supplementary material including more detailed methods, and supplementary tables and figures for "A Reduced Digital Motor Assessment using Multimodal Biomarkers in Parkinson’s Disease"

#### 1. Supplementary Methods

##### | 1.1 Surface EMG recording and electrode placement

Two sensors were used for all tasks except pronation–supination (T3), which was recorded with a single sensor, and were repositioned between tasks. Recordings were made on the more-affected side in participants with Parkinson's disease (PD) and on the dominant side in healthy controls (HC), reflecting the asymmetric onset of parkinsonian motor signs [1]. Skin preparation followed Surface Electromyography (EMG) for the Non-Invasive Assessment of Muscles (SENIAM) recommendations [2].

**TABLE S1** Surface EMG electrode placement per motor task. Target muscles for each of the seven Movement Disorder Society–Unified Parkinson's Disease Rating Scale Part III (MDS-UPDRS III) tasks.

| Task | Motor task (MDS-UPDRS III item) | Target muscle(s) |
|---|---|---|
| T1 | Finger tapping (3.4) | Flexor digitorum superficialis (FDS), Extensor digitorum (ED) |
| T2 | Hand movements (3.5, hand opening–closing) | FDS, ED |
| T3 | Pronation–supination (3.6) | Pronator teres (PT) |
| T4 | Postural tremor, arms extended (3.15) | Flexor carpi radialis (FCR), Extensor carpi radialis (ECR) |
| T5 | Toe tapping (3.7) | Tibialis anterior (TA), Gastrocnemius (GAS) |
| T6 | Gait, 10 m walk-and-turn (3.10) | TA, GAS |
| T7 | Leg agility (3.8, foot stomping) | Rectus femoris (RF), Semitendinosus (ST) |

##### | 1.2 EMG signal preprocessing

Surface EMG was recorded at 1000 Hz. Each channel was mean-centred, band-pass filtered (1–450 Hz, fourth-order Butterworth) and notch filtered (50 Hz, second-order IIR notch, $Q = 30$), both applied with zero-phase (forward–backward) filtering. The linear envelope was obtained by full-wave rectification followed by a fourth-order 10 Hz low-pass Butterworth filter. Three representations were retained: the filtered signal, its full-wave rectified version, and the 10-Hz low-pass linear envelope.

**Normalization.** For the repetitive and gait tasks, each muscle was amplitude-normalized to the 95th percentile of its linear envelope. Amplitude normalization was applied to the filtered signal, from which the rectified signal and envelope were then recomputed, so that all three representations share a single scaling constant. The postural task (T4) was retained in absolute millivolts.

**Burst segmentation.** For the repetitive and gait tasks, activation bursts were delineated from the linear envelope by a two-pass peak-detection procedure: The height of a genuine burst varies between participants and muscles, so the detection threshold cannot be fixed in advance. A first pass therefore detected peaks above the 70th percentile of the envelope – samples that lie within clearly active periods regardless of the recording's overall amplitude – and the median height of these peaks was taken as an estimate of the typical burst amplitude for that recording. The working threshold was then set to 0.3 times this median, low enough to also capture smaller genuine bursts missed by the first pass. Each burst's onset and offset were placed at the surrounding envelope minima. Relaxation periods were defined as the intervals between consecutive bursts (offset → next onset). Burst-level and relaxation-level features were summarized across repetitions by their mean, standard deviation and coefficient of variation.

**Feature computation.** Amplitude features were computed as root-mean-square ($\sqrt{\overline{x^2}}$), mean absolute value ($\overline{|x|}$), integrated EMG ($\sum |x|/f_s$), waveform length ($\sum |\Delta x|$), and zero-crossing rate. Spectral features were derived from the power spectral density (Welch's method, 2-s segments, `nperseg` $= \min(N, 2f_s)$) of the filtered signal and the envelope. Signal-complexity features used the `antropy` library (v0.1.9): Higuchi fractal dimension [3], sample entropy [4], permutation entropy [5], and detrended fluctuation analysis; the DFA exponent [6].

**Frequency bands.** Spectral features used low (20–40 Hz), mid (40–150 Hz), and high (150–450 Hz) sub-bands; a tremor band (3.5–7.5 Hz), selected to encompass the characteristic frequencies of parkinsonian rest and postural tremor [7]. Tremor features included the tremor ratio (band power / total power) and tremor peak frequency (frequency of maximal power within the tremor band). The envelope fluctuation rate was defined as the number of envelope-slope reversals per second.

#### 1.3 EMG features

**TABLE S2** EMG feature dictionary. Features are computed per muscle and prefixed by muscle (e.g. `fcr_`, `ecr_`) or, for cross-muscle features, `inter_`. **Panel A**: postural task (T4), single continuous epoch, kept in absolute mV (no normalisation). **Panel B**: repetitive and gait tasks (T1–T3, T5–T7), full active segment plus per-cycle bursts (per-burst features summarized across bursts by mean, SD and CV, where CV=SD/mean). Three signal representations are used throughout: the band-pass *filtered* signal (1–450 Hz, signed), the *rectified* signal (|filtered|), and the linear *envelope* (rectified, low-pass 10 Hz). Bands as in Supplementary Section 1.2.

| Feature | Signal | Definition |
|---|---|---|
| **Panel A — Postural task (T4)** | | |
| *Filtered signal* | | |
| RMS, mean absolute value, variance | Filtered | Amplitude measures |
| Zero-crossing rate | Filtered | Rate of sign changes |
| Skewness, kurtosis | Filtered | Amplitude-distribution shape |
| Sample entropy | Filtered | Regularity of the amplitude series |
| Higuchi fractal dimension | Filtered | Waveform complexity |
| Permutation entropy | Filtered | Ordinal-pattern complexity |
| DFA $\alpha$ (signal) | Filtered | Detrended-fluctuation scaling exponent of the filtered signal |
| Mean, median frequency | Filtered | Power-weighted mean; spectral median |
| Band power (20–40 / 40–150 / 150–450 Hz) | Filtered | Integrated power in each motor-unit sub-band |
| *Envelope-derived complexity* | | |
| DFA $\alpha$ (envelope) | Envelope | DFA scaling exponent of the activation envelope [a] |
| *Rectified signal* | | |
| Integrated EMG, waveform length | Rectified | Area and cumulative path length |

*continued*

| Feature | Signal | Definition |
|---|---|---|
| Mean absolute value (rectified) | Rectified | Mean rectified amplitude |
| Peak amplitude | Rectified | Maximum rectified amplitude |
| Turns rate | Rectified | Slope reversals per second |
| *Envelope* | | |
| Envelope mean, SD, CV, range | Envelope | Linear-envelope amplitude statistics |
| Activity flag | Envelope | Presence of supra-threshold activity |
| Tremor power, total power, tremor ratio | Envelope | Power in 3.5–7.5 Hz (absolute; and relative to 0–20 Hz) |
| Tremor peak frequency | Envelope | Frequency of peak power in the tremor band |
| Dominant frequency, dominant-peak ratio | Envelope | Peak frequency (0–12 Hz); peak-to-mean spectral sharpness |
| Spectral entropy | Envelope | Entropy of the normalised envelope spectrum |
| Envelope fluctuation rate | Envelope | Envelope slope reversals per second |
| Windowed tremor stability | Envelope | Mean and SD across 2 s windows of tremor ratio, tremor peak frequency, dominant frequency, and envelope sample entropy |
| *Inter-muscle (FCR–ECR)* | | |
| Co-contraction index [a] | Envelope | Antagonist co-activation |
| Envelope correlation | Envelope | Pearson correlation of the two envelopes |
| RMS ratio | Filtered | RMS(FCR) / RMS(ECR) |
| Tremor coherence | Envelope | Envelope coherence in 3.5–7.5 Hz |
| **Panel B — Repetitive and gait tasks (T1–T3, T5–T7)** | | |
| *Full active segment — filtered* | | |
| RMS | Filtered | Root-mean-square amplitude |
| Sample entropy, Higuchi fractal dimension, permutation entropy | Filtered | Complexity measures |
| Skewness, kurtosis | Filtered | Amplitude-distribution shape |
| Mean, median frequency | Filtered | Spectral centroid; spectral median |
| Band power (20–40 / 40–150 / 150–450 Hz) | Filtered | Motor-unit sub-band powers |
| *Full active segment — envelope* | | |
| DFA $\alpha$ | Envelope | DFA scaling exponent of the envelope (gait/movement amplitude rhythm) |
| Tremor ratio, tremor peak frequency | Envelope | Tremor-band (3.5–7.5 Hz) content |
| Total power | Envelope | Envelope power, 0–20 Hz |
| Dominant frequency, dominant-peak ratio | Envelope | Peak envelope frequency; peak-to-mean sharpness |
| Spectral entropy | Envelope | Entropy of the envelope spectrum |
| Envelope mean, SD | Envelope | Envelope amplitude statistics |

*continued*

| Feature | Signal | Definition |
|---|---|---|
| Envelope fluctuation rate | Envelope | Envelope slope reversals per second |
| *Full active segment — rectified* | | |
| Integrated EMG, waveform length | Rectified | Area and cumulative path length |
| Turns rate | Rectified | Slope reversals per second |
| *Burst statistics (across bursts: mean / SD / CV)* | | |
| Cadence (bursts $s^{-1}$) | Env. bursts | Burst rate over the active segment |
| Inter-burst interval (mean, CV) | Env. bursts | Peak-to-peak spacing and its variability |
| Burst peak amplitude (mean, SD) | Env. bursts | Per-burst peak amplitude |
| Amplitude decline slope, halves ratio | Env. bursts | Trend of burst amplitude across the trial |
| Burst duration (mean, SD, CV) | Env. bursts | Burst timing |
| Rise, fall time (mean, SD) | Env. bursts | Onset-to-peak and peak-to-offset times |
| Burst area | Env. bursts | iEMG per burst |
| Duty cycle | Env. bursts | Fraction of active time spent bursting |
| *Per-burst segment features (across bursts: mean / SD / CV)* | | |
| Zero-crossings, variance | Filtered bursts | Per-burst time-domain measures |
| Tremor power, tremor ratio, tremor and dominant peak frequency | Filtered bursts | Per-burst spectral content |
| RMS, iEMG, peak, waveform length | Rectified bursts | Per-burst amplitude measures |
| *Relaxation statistics (between bursts)* | | |
| Relaxation duration (mean, SD, CV) | Envelope | Inter-burst interval length |
| Relaxation amplitude (mean, SD), RMS | Envelope | Background activation between bursts |
| Relaxation-to-burst ratio | Envelope | Inter-burst vs burst amplitude |
| Rest-tremor power, tremor ratio, dominant frequency (mean, SD) | Filtered | Tremor content of the inter-burst periods |
| *Inter-muscle (two-muscle tasks only)* [b] | | |
| Co-contraction index [a] | Envelope | Antagonist co-activation over active windows |
| Envelope correlation (global; per-burst mean, SD) | Envelope | Trial-wide and local envelope correlation |
| RMS ratio | Filtered | RMS ratio of the two muscles |
| Cadence coherence | Envelope | Envelope coherence in the cadence band |
| Phase lag (mean, SD, $\lvert\cdot\rvert$ mean) | Envelope | Burst-peak timing offset between muscles (ms) |

*Abbreviations:* CV, coefficient of variation; DFA, detrended fluctuation analysis; ECR, extensor carpi radialis; EMG, electromyography; FCR, flexor carpi radialis; iEMG, integrated EMG; RMS, root-mean-square; SD, standard deviation. [a] Co-contraction index [8]: $\overline{2\min(a,b)/(a+b)}$, the sample-wise mean of the normalized overlap between the two antagonist envelopes (0 = no co-activation, 1 = identical activation). [b] Inter-muscle features apply to the two-muscle tasks (all except T3).

#### | 1.4 Musculoskeletal modeling and features

Whole-body kinematics were exported as skeletal motion .bvh (BioVision Hierarchy) files from the Movella Xsens MVN software, and used to drive a subject-scaled full-body musculoskeletal (MSK) model in the AnyBody Modeling System [9, 10] (via AnyPyTools v1.20.1), personalized by independent (per-axis) anthropometric scaling and by each participant's body mass, height, and motion-capture–derived segment lengths, and solved by inverse kinematics (marker tracking) followed by inverse dynamics, with ground-reaction forces predicted.

**Normalization.** Ground reaction forces, joint reaction forces and muscle forces were normalized by body weight in Newtons ($BW = m \cdot g$) [11, 12], and joint moments by $BW\times$ height [13]. Stride and step length were additionally expressed relative to standing height [14, 15]. Because arm-swing range of motion scales with walking speed and gait was performed at a self-selected pace, arm-swing range of motion (ROM) was additionally normalized by gait speed [16, 17]. Joint angles and angular velocities are size-independent and were left unnormalized; joint angles and joint angular velocities are reported in degrees and degrees $s^{-1}$, and segment angular-velocity magnitudes (hand, forearm) in rad $s^{-1}$.

**TABLE S3 Anatomical scope of the MSK model per task.** Joints, muscles (activation and force), and end-point/gait measures from which features were extracted. The feature families derived from these signals are listed in Table S4.

| Task | Joints | Muscles | End-point / gait |
|---|---|---|---|
| T1 Finger tapping | Shoulder, elbow, wrist | BIC, TRI, FCR, FCU, ECRB, ECU, ED, FDS, FDP | Hand, forearm |
| T2 Hand movements | Shoulder, elbow, wrist | As T1 | Hand, forearm |
| T3 Pron–supination | Shoulder, elbow, wrist | BIC, TRI, FCR, FCU, ECRB, ECU, PT, PQ, SUP | Hand, forearm |
| T4 Postural tremor | Shoulder, elbow, wrist | BIC, TRI, FCR, ECRB | Hand, forearm |
| T5 Toe tapping | Hip, knee, ankle, subtalar | TA, MG, SOL, TP, PL | Foot, GRF |
| T6 Gait (walk+turn) | Hip, knee, ankle, subtalar, pelvis, trunk; shoulder, elbow | TA, MG, LG, SOL, PL, VL, GMED | Foot, arm swing, stride/step, cadence, speed, turn, GRF |
| T7 Leg agility | Hip, knee, ankle, subtalar | RF, ST, BF, SART | Foot, GRF |

*Abbreviations:* BF, biceps femoris; BIC, biceps brachii; ECRB, extensor carpi radialis brevis; ECU, extensor carpi ulnaris; ED, extensor digitorum; FCR, flexor carpi radialis; FCU, flexor carpi ulnaris; FDP, flexor digitorum profundus; FDS, flexor digitorum superficialis; GMED, gluteus medius; GRF, ground reaction force; LG, lateral gastrocnemius; MG, medial gastrocnemius; PL, peroneus longus; PQ, pronator quadratus; PT, pronator teres; RF, rectus femoris; SART, sartorius; SOL, soleus; ST, semitendinosus; SUP, supinator; TA, tibialis anterior; TP, tibialis posterior; TRI, triceps brachii; VL, vastus lateralis.

**TABLE S4** Musculoskeletal feature families, source signal, applicable tasks (Upper = T1–T4; Lower = T5–T7), and definition. Features are computed per joint degree of freedom and per muscle on the recorded side; the joints, muscles and end-point measures to which each family was applied are listed per task in Table S3.

| Feature | Source signal | Tasks | Definition |
|---|---|---|---|
| Angle mean, SD, drift | Joint angle | Upper, Lower | Level, variability, linear trend over the trial |
| Range of motion | Joint angle | Upper, Lower | Angular excursion of the joint |
| Tremor ratio, tremor peak frequency | Joint angle, angular velocity, activation | Upper, Lower | Tremor-band content of the signal |
| Angular velocity RMS | Angular velocity | Upper, Lower | Root-mean-square angular velocity |
| Angular velocity spectral / sample entropy | Angular velocity | Upper, Lower | Spectral and sample-entropy irregularity |
| Movement smoothness (SPARC) | Angular velocity | Upper, Lower | Spectral arc length; log dimensionless jerk |
| Hand speed (RMS, dominant freq., spectral entropy, drift) | Hand end-point | Upper | Speed magnitude, spectral peak, irregularity, trend |
| Hand smoothness (SPARC) | Hand end-point | Upper | Smoothness of the hand-speed profile |
| Hand position (SD, range, drift) | Hand end-point | Upper | Per-axis positional variability, excursion, trend |
| Muscle activation (mean, SD, drift, skewness, kurtosis) | Simulated activation | Upper, Lower | Level, variability, trend, distribution shape |
| Activation spectral (dominant freq., tremor ratio, spectral entropy) | Simulated activation | Upper | Frequency content of forearm-muscle activation |
| Muscle force (mean, skewness, kurtosis) | Simulated force | Upper, Lower | Level and distribution shape (BW-normalized) |
| Co-contraction index (activation, force) [a] | Activation/force pair | Upper, Lower | Antagonist co-activation at a joint |
| Joint moment (mean, SD, tremor ratio, tremor peak freq.) | Inverse-dynamics moment | Upper, Lower | Net joint moment (BW×height normalized) |
| Joint reaction force | Reaction force | Upper, Lower | Internal joint contact force (BW-normalized); not available for T4 |
| Ground reaction force | Predicted GRF | Lower | Foot–ground force magnitude and peaks |
| Spatiotemporal (stride, step, cadence, speed, stance, turn) | Segment kinematics | Lower | Gait timing and distance measures |
| Trunk/pelvis (flexion, sway, rotation), arm swing | Segment kinematics | Lower | Axial posture and arm-swing amplitude |
| Bilateral asymmetry index [b] | Recorded vs contralateral | Upper, Lower | Normalized side-to-side difference |

[a] Co-contraction index (CCI), adapted from Falconer and Winter [8]: $\overline{2\min(a,b)/(a+b)}$, the sample-wise mean of the normalised overlap

between the two antagonist signals (0 = no co-activation, 1 = identical). Computed from antagonist muscle activations and, in parallel, forces spanning a joint (pairs: wrist flexors–extensors, T1/T2; pronator teres–supinator, biceps–pronator teres and pronator quadratus–supinator, T3; wrist and elbow, T4; gastrocnemius–tibialis anterior, T5/T6; rectus femoris–semitendinosus, T7). [b]Bilateral asymmetry index, adapted from [18, 19]: $(x_{\text{rec}} - x_{\text{con}})/\max(|x_{\text{rec}}|, |x_{\text{con}}|)$, where $x_{\text{rec}}$ is the value on the recorded side (more-affected side in PD, dominant side in controls) and $x_{\text{con}}$ the contralateral side; positive values indicate a larger value on the recorded side. Available for the MSK modality only, as EMG was recorded unilaterally.

*Abbreviations:* BW, body weight; RMS, root-mean-square; SD, standard deviation; SPARC, spectral arc length.

#### | 1.5 Classification pipeline and validation

**Feature selection (within each training fold, fixed order).** Median imputation; near-zero-variance removal (variance threshold 0.01); a supervised correlation filter that, for each pair with $|r| > 0.80$, discarded the feature less correlated with the group label; mutual-information (MI) ranking against the group label [20, 21], which captures both linear and non-linear dependence, retaining features up to the knee of the ranked MI curve. The knee was identified using the maximum-deviation criterion [22], defined as the point of greatest vertical gap between the curve and the straight line joining its first and last points. The resulting feature subset was bounded to 5–100 features; standardization; and embedded $\ell_1$-penalized logistic regression (LASSO, $C = 0.5$), reducing the pool to a final subset of three to five features. The cap keeps the number of features modest relative to the sample size, limiting overfitting in a penalized model [23]. No step used the test fold [24].

**Classifier panel.** Eleven classifiers were evaluated per cell: logistic regression (elastic-net penalty, $C = 1.0$, $\ell_1$ ratio 0.5), linear, polynomial and RBF support-vector machines, Gaussian naive Bayes, $k$-nearest neighbours, decision tree, random forest, gradient boosting, XGBoost, linear discriminant analysis (Ledoit–Wolf shrinkage). All used scikit-learn default hyperparameters; none were tuned.

**Implementation of the validation.** For each task–configuration cell the classifiers were compared under a single stratified 10-fold cross-validation (`random_state`=42), and the classifier with the highest balanced accuracy was retained for all subsequent analyses. The retained classifier was re-evaluated over 30 stratified 10-fold partitions generated with seeds 42–71, giving 300 train–test folds per cell. Metrics were computed at a decision threshold of 0.5. Point estimates are the pooled out-of-fold predictions; performance across repetitions is summarized as the mean $\pm 1.96\,\text{SD}$, which describes the 95% interval variability of the estimate across partitions.

The permutation $p$-value was computed as $p = (1 + \#\{\text{perm} \geq \text{observed}\})/(1 + 500)$, where *observed* is the performance of the model trained on the true labels, *perm* that of a model trained on permuted labels [25, 26].

#### | 1.6 Multimodal feature configuration

In the multimodal configuration, the filtering, MI ranking and knee-point steps were applied independently to the EMG and MSK feature sets within each training fold; the two candidate pools were then concatenated and passed to a single LASSO step, so the final 3–5 features could be drawn from either modality. A joint ranking would let the larger MSK set dominate the pool before the embedded step.

#### | 1.7 Modality and task fusion

The meta-model was an $\ell_1$-penalized logistic regression (`liblinear`, $C = 1.0$, balanced class weights) fitted on the $61 \times 7$ matrix of task-specific out-of-fold PD probabilities, and evaluated in its own stratified 10-fold cross-validation (`random_state`=42).

#### 1.8 Task complementarity

Task contributions were the mean meta-model coefficient across the meta-model cross-validation folds, with each task's fold selection frequency. Redundancy was quantified between the columns of the $61 \times 7$ matrix as the pairwise Pearson correlation and MI.

#### 1.9 Stable-feature identification

Selection stability was quantified as the proportion of cross-validation folds in which each feature was retained within its own modality. Features selected in more than 50% of folds were considered stable [27, 28]. Stable features were retained as candidate biomarkers if they also differed significantly between groups (Mann–Whitney U, $p < 0.05$, uncorrected). Selection frequency in the multimodal configuration was additionally reported to characterize whether each feature remained selected when competing with features from the other modality.

### 2. Supplementary results

**TABLE S5 Sensitivity and specificity per motor task and modality.** Pooled cross-validated estimates: each subject's out-of-fold prediction is collected across the ten folds and the metrics computed once on the complete set, at a threshold of 0.5. The best-performing classifier in each task–modality combination was selected by balanced accuracy.

| | EMG | | Musculoskeletal | | Multimodal | |
|---|---|---|---|---|---|---|
| Task | Sens. | Spec. | Sens. | Spec. | Sens. | Spec. |
| T1 Finger tapping | 0.65 | 0.63 | 0.81 | 0.73 | 0.84 | 0.80 |
| T2 Hand movements | 0.84 | 0.94 | 0.65 | 0.83 | 0.84 | 0.77 |
| T3 Pron–supination | 0.81 | 0.90 | 0.84 | 0.77 | 0.87 | 0.95 |
| T4 Postural tremor | 0.84 | 0.73 | 0.71 | 0.80 | 0.74 | 0.73 |
| T5 Toe tapping | 0.71 | 0.60 | 0.52 | 0.60 | 0.58 | 0.70 |
| T6 Gait | 0.55 | 0.57 | 0.71 | 0.60 | 0.71 | 0.60 |
| T7 Leg agility | 0.65 | 0.63 | 0.65 | 0.63 | 0.55 | 0.63 |

*Abbreviations:* EMG, surface electromyography; Sens., sensitivity; Spec., specificity.

**TABLE S6 Task contribution across the three fusion variants.** Mean LASSO meta-model coefficient ± SD across cross-validation folds, with the fraction of folds in which each task was selected in parentheses. Tasks with a coefficient of zero were shrunk out of the model in all folds.

| Task | EMG fusion | MSK fusion | Multimodal fusion |
|---|---|---|---|
| T1 Finger tapping | 0 | +1.91 ± 0.26 (1.0) | +0.38 ± 0.41 (0.9) |
| T2 Hand movements | +1.57 ± 0.34 (1.0) | +0.94 ± 0.30 (1.0) | +0.82 ± 0.50 (1.0) |
| T3 Pron–supination | +2.78 ± 0.21 (1.0) | +1.97 ± 0.29 (1.0) | +3.76 ± 0.30 (1.0) |
| T4 Postural tremor | +0.94 ± 0.29 (1.0) | +0.83 ± 0.27 (1.0) | +0.94 ± 0.18 (1.0) |
| T5 Toe tapping | 0 | 0 | 0 |
| T6 Gait | 0 | 0 | 0 |
| T7 Leg agility | 0 | 0 | 0 |

*Abbreviations:* EMG, electromyography; MSK, musculoskeletal; LASSO, least absolute shrinkage and selection operator; SD, standard deviation.

**TABLE S7 Association of the fused prediction with clinical and demographic variables.** Spearman rank correlation between the fused Parkinson's disease probability and each variable, computed within each group separately (PD $n$ = 31, HC $n$ = 30). Clinical variables are recorded for the PD group only. For sex, the statistic is the common-language effect size (the probability that a randomly chosen woman scores higher than a randomly chosen man), with a Mann–Whitney $p$-value.

| | | PD | | HC | |
|---|---|---|---|---|---|
| | Variable | $\rho$ | $p$ | $\rho$ | $p$ |
| *Clinical* | Hoehn & Yahr stage | +0.10 | 0.58 | – | – |
| | Time since last dose | +0.25 | 0.17 | – | – |
| *Demographic* | Age | +0.28 | 0.12 | −0.13 | 0.49 |
| | BMI | −0.06 | 0.73 | +0.26 | 0.17 |
| | Sex (F vs M) | 0.58 | 0.49 | 0.50 | 0.98 |

*Abbreviations:* BMI, body mass index; HC, healthy controls; PD, Parkinson's disease.

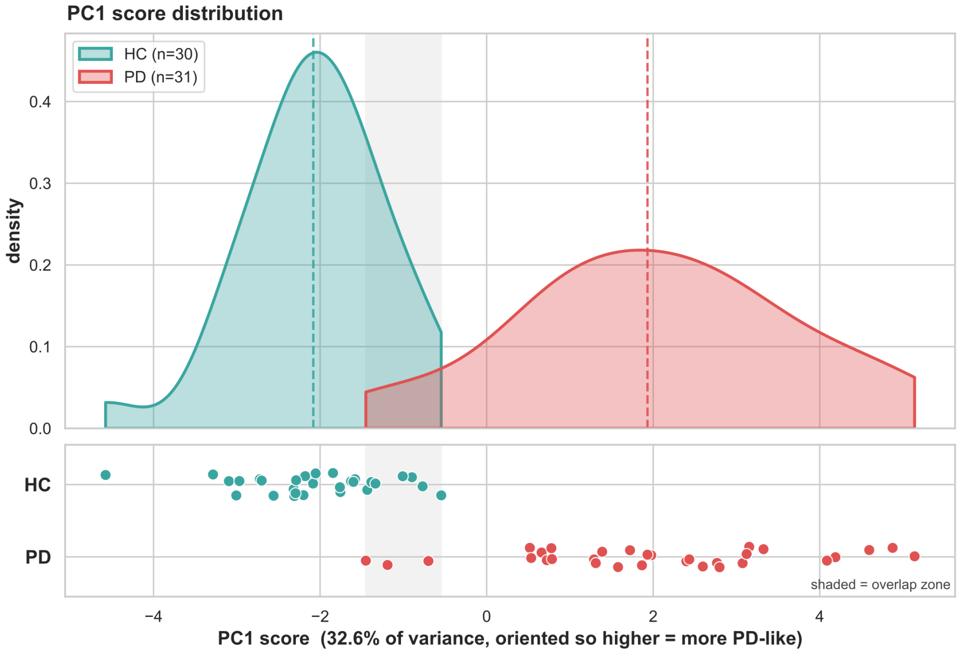


**FIGURE S1** **Distribution of PC1 scores.** Kernel density estimates and individual scores for healthy controls (HC, n = 30) and patients with Parkinson's disease (PD, n = 31) along the first principal component of the stable biomarker set, accounting for 32.6% of variance. Dashed lines mark group means.

**TABLE S8** **Feature-selection stability of the 18 retained biomarkers.** Selection rate is the fraction of cross-validation folds in which LASSO retained each feature, over 30 repeats of 10-fold cross-validation (300 folds). *Modality* is the rate within the feature's own modality; *Fusion* is the rate in the multimodal model, where EMG and MSK features compete for the same 3–5 selection slots. Entries listed as T1, T2 were stable in both tasks; the measurement from the more significant task is reported.

| # | Biomarker | Task | Modality | Selection rate | |
|---|---|---|---|---|---|
| | | | | Modality | Fusion |
| 1 | Pronator teres – env. fluctuation rate | T3 | EMG | 1.00 | 1.00 |
| 2 | Pronator teres – relaxation-duration CV | T3 | sEMG | 0.99 | 0.82 |
| 3 | Biceps–pronator teres co-contraction | T3 | MSK | 0.57 | 0.01 |
| 4 | Hand – angular-velocity peak | T3 | MSK | 0.95 | 0.39 |
| 5 | Wrist – flexion moment RMS asymmetry | T3 | MSK | 0.86 | 0.54 |
| 6 | Pronator teres – per-burst dominant freq. | T3 | EMG | 0.57 | 0.08 |
| 7 | ECR – mean dominant frequency | T4 | EMG | 0.99 | 0.82 |
| 8 | ECR – tremor ratio | T4 | EMG | 0.75 | 0.05 |
| 9 | Elbow – flexion angular-velocity RMS | T4 | MSK | 0.67 | 0.28 |
| 10 | FCR – envelope DFA $\alpha$ | T4 | EMG | 0.53 | 0.22 |
| 11 | FDS – env. fluctuation rate | T2 | EMG | 0.91 | 0.94 |
| 12 | FDS – per-burst peak amplitude SD | T2 | EMG | 0.89 | 0.94 |
| 13 | Wrist – flexion velocity kurtosis | T2 | MSK | 0.91 | 0.25 |
| 14 | Elbow – mean flexion angle | T2 | MSK | 0.59 | 0.01 |
| 15 | Forearm – angular-velocity RMS | T1 | MSK | 0.55 | 0.72 |
| 16 | Shoulder – flexion angle drift | T1, T2 | MSK | 0.99 | 0.94 |
| 17 | ED – relaxation dominant freq. SD | T1, T2 | EMG | 0.66 | 0.80 |
| 18 | FDS – tremor ratio | T1 | EMG | 0.50[a] | 0.02 |

[a] Rounded from 0.503; the inclusion threshold was applied to unrounded rates.

*Abbreviations:* CV, coefficient of variation; DFA, detrended fluctuation analysis; ECR, extensor carpi radialis; ED, extensor digitorum; EMG, electromyography; env., envelope; FCR, flexor carpi radialis; FDS, flexor digitorum superficialis; freq., frequency; LASSO, least absolute shrinkage and selection operator; MSK, musculoskeletal; RMS, root-mean-square; SD, standard deviation.

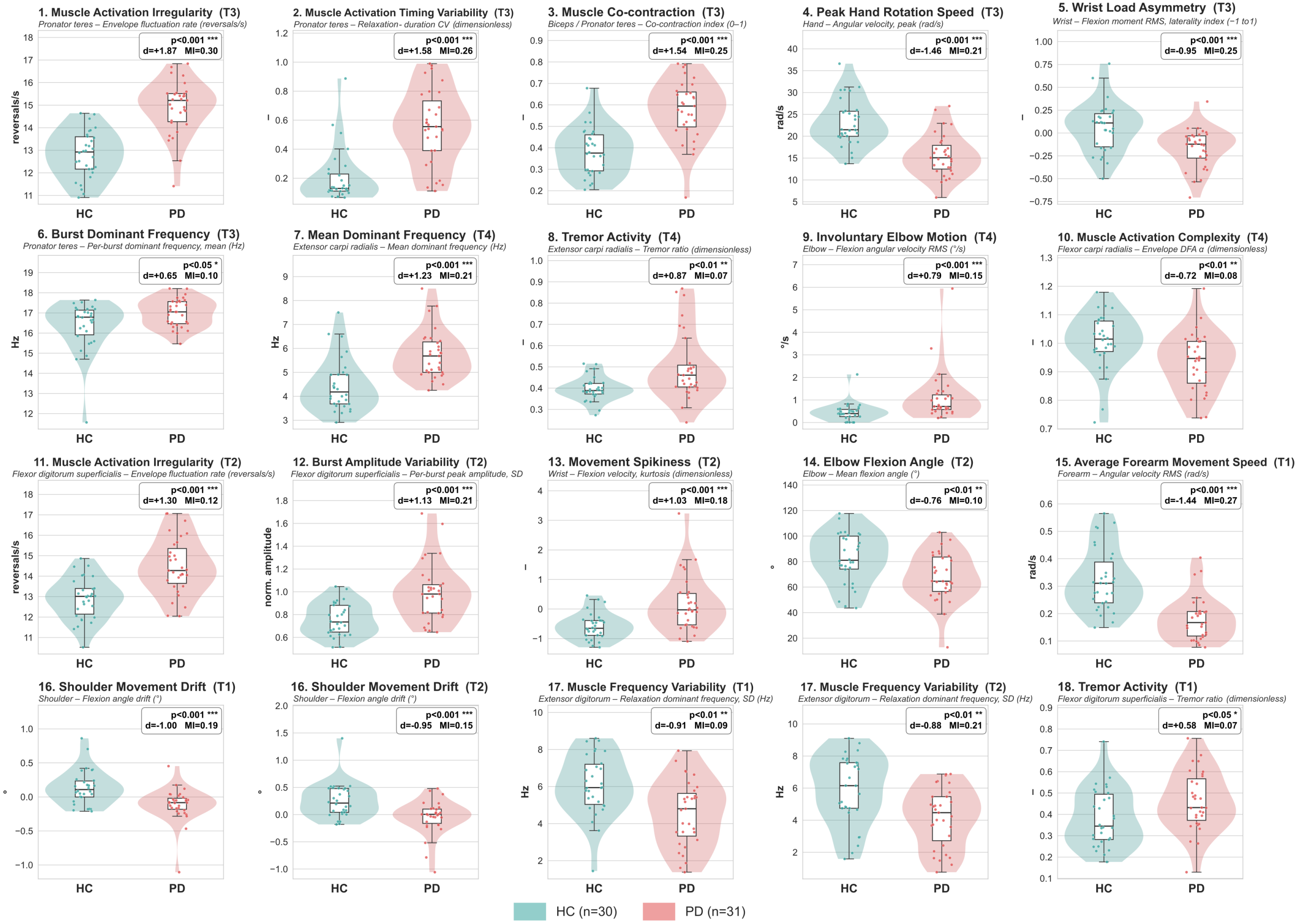


**FIGURE S2 Group distributions of the 18 stable digital biomarkers.** Combined box-and-violin plots comparing healthy controls (HC, $n$ = 30) and Parkinson's disease patients (PD, $n$ = 31) ordered as Fig. 3A. Boxes show the median and interquartile range; overlaid points are individual subjects. Each panel is annotated with the effect size (Cohen's $d$), MI with group (MI), and Mann–Whitney U-test significance ($^{***}p < 0.001$, $^{**}p < 0.01$, $^{*}p < 0.05$). Units follow each feature's physical scale.
